# Factors associated with transmission in COVID-19 outbreaks in long-term care facilities

**DOI:** 10.1101/2021.07.12.21260345

**Authors:** Rohit Vijh, Carmen H. Ng, Mehdi Shirmaleki, Aamir Bharmal

**Author notes:** **Corresponding Author:** Aamir Bharmal, MD MPH, Office of the Medical Health Officer, Fraser Health, Suite 400, Central City Tower, 13450 – 102 Avenue, Surrey, BC V3T 0H1. **Funding Sources**: This study did not receive any funding from agencies in the public, commercial, or not-for-profit sectors.

## Abstract

**Background:** Severe acute respiratory syndrome coronavirus 2 (SARS-CoV-2) has had a disproportionate impact on residents in long-term care facilities (LTCFs). Through our experience and data from managing COVID-19 exposures and outbreaks in LTCFs in the Fraser Health region in British Columbia, Canada, we identified risk factors associated with outbreak severity to inform current outbreak management strategies and future pandemic preparedness planning efforts.

**Methods:** We used a retrospective cohort study design to evaluate the association between non-modifiable factors (facility building, organization level, and resident population characteristics), modifiable factors (assessments for infection prevention and control (IPC) and public health measures), and severity of COVID-19 outbreaks (attack rate) in LTCFs. We modelled the COVID-19 attack rates in LTCF outbreaks using negative binomial regression models.

**Results:** From March 1, 2020 to January 10, 2021, a total of 145 exposures to at least one confirmed case of COVID-19 in 82 LTCFs occurred. For every item not met in the assessment tool, a 22% increase in the attack rate was observed (rate ratio 1.2 [95% CI 1.1 – 1.4]) after adjusting for other risk factors such as age of the facility, index case type (resident vs. staff) and proportion of single bed rooms.

**Conclusion:** Our findings highlight the importance of assessing IPC and public health measures for outbreak management. They also demonstrate the important modifiable and non-modifiable risk factors associated with COVID-19 outbreaks in our jurisdiction. We hope these findings will inform ongoing outbreak management and future pandemic planning efforts.

## Introduction

Severe acute respiratory syndrome coronavirus 2 (SARS-CoV-2) has had a disproportionate impact on vulnerable populations, particularly residents in long-term care facilities (LTCFs). The proportion of COVID-19 deaths from LTCFs in Canada has been higher compared to other countries.^1^ Between March 1, 2020 to February 15, 2021, around 70% of deaths from SARS-CoV-2 in the province of British Columbia, Canada, were among LTCF residents.^2^ A considerable amount of evidence has emerged to understand potential risk factors for COVID-19 outbreaks and spread.^3–12^ Furthermore, evidence has also emerged on useful strategies to contain and prevent the spread of infection in LTCFs.^13–16^

Previous studies in Canada examining risk factors of COVID-19 outbreaks in LTCFs have focused on resident level and organizational level factors only. Several other risk factors such as building design, community incidence (facilitating frequent introductions), for-profit status, rating status and resident population characteristics, such as the proportion with dementia, have been identified to increase both the risk of an outbreak occurring at LTCFs and its severity.^3,6,7^ A number of these risk factors are challenging to address and not within the control of LTCF operators. Therefore, infection prevention and control (IPC) best practices are critical to employ to reduce transmission and to prevent outbreaks.

Standardized audit tools which assess the implementation of IPC measures have been demonstrated to reduce communicable disease transmission in LTCFs^17,18^. Near the beginning of the COVID-19 pandemic, a standardized outbreak prevention assessment tool was created and deployed to LTCFs in the Fraser Health region in British Columbia, Canada at the start of an outbreak in order to prevent further transmission. The tool identifies areas of weakness in IPC practices, public health measures and pandemic preparedness and requires LTCF operators to respond immediately to mitigate identified areas of concern for further transmission. As the majority of the COVID-19 outbreaks in LTCFs in the province of British Columbia occurred in the Fraser Health region, a robust examination of a large sample of LTCF outbreaks was done to evaluate the tool and identify both modifiable and non-modifiable risk factors associated with COVID-19 transmission within LTCFs.

The objective of our study was to identify risk factors associated with LTCF outbreak attack rates (as a measure for severity) to inform both current outbreak management strategies in LTCFs and future pandemic preparedness planning efforts. Factors examined included (1) building characteristics, (2) organization level characteristics, (3) resident population characteristics, (4) IPC and public health measures based on an assessment during the outbreak, (5) the community COVID-19 incidence rate, and (6) the index case being a resident or a staff member.

## Methods

### Study Design

We utilized a retrospective cohort design to evaluate the association between non-modifiable factors (facility building, organization level, and resident population characteristics), modifiable factors (assessments for IPC and public health measures), and severity of COVID-19 outbreaks in LTCFs. This analysis was exempt from ethics review as it was a public health surveillance and quality improvement activity in response to the COVID-19 emergency response.

Our analysis was divided into two steps. The first was a descriptive summary of LTCF building, organization level and resident population characteristics to inform variables to include in the regression analysis. The second step was to use a regression analysis to identify factors associated with outbreak severity while controlling for other factors. Predictors included in the regression analysis included factors identified from the descriptive analysis, scores from the assessment tool, and other confounding variables identified from the literature.

### Study Setting

Responsibilities for health care services in the province of British Columbia, Canada, are organized into a Provincial Health Services Authority, five geographic regional health authorities (RHAs), and a First Nations Health Authority.^19^ Provincially, 33% of publicly-funded LTC beds are operated by RHAs, 32% are operated by not-for-profit societies contracted by RHAs, and 35% are operated by for-profit businesses (private pay LTCFs).^20^

Fraser Health is a RHA serving more than 1.9 million people (approximately 38% of the province’s population) across a mixed urban and rural geographic area straddling 20 different communities, including approximately 62,000 Indigenous Peoples. The region has had the highest burden of COVID-19 cases and LTCF outbreaks in the province. Fraser Health operates 12 acute care hospitals, and operates or is affiliated with around 8,400 beds in 82 LTCFs^21^. Almost 80% of the publicly-funded LTCF beds are operated by affiliated providers and the remainder are in health authority owned and operated LTCFs. There were 12 private pay LTCFs at the time of this study.

In the event of a COVID-19 exposure or outbreak, the health authority supports all LTCFs in the region regardless of funding structure. Fraser Health provides standardized protocols for COVID-19 exposure and outbreak management. At the onset of monitoring for a COVID-19 exposure or a declared outbreak, a standardized assessment is done to assess facility readiness for COVID-19 outbreaks and to inform outbreak management by identifying areas of weakness in IPC practices and public health measures. Additional details regarding outbreak management and the assessment tool can be found in Appendix A.

A LTCF is considered to have a COVID-19 exposure when a lab-confirmed COVID-19 case^22^ among staff, residents, or another individual who was at the LTCF during their infectious period is identified, and there is risk of exposure to other individuals on-site. Up until November 9, 2020, an outbreak was declared whenever there was a COVID-19 exposure at a LTCF. From November 9 onwards, an outbreak was declared only when there was suspected or confirmed transmission within the LTCF. LTCFs being monitored due to a COVID-19 exposure are still required to maintain IPC practices and protocols similar to when an outbreak is declared, including isolation of exposed residents/staff, testing, and cessation of group activities, social visits, and admissions and transfers of residents.

### Study Population (Inclusion Criteria)

All LTCFs in the region (n=93 unique facilities) were considered for inclusion in the analysis. Private pay LTCFs (n=12), a new LTCF built in April 2020 (n=1) and a pediatric LTCF (n=1) were excluded from the descriptive analysis as building, organization level and resident population characteristics were only available for publicly funded or Fraser Health owned adult LTCFs in operation during April 1, 2019 to March 31, 2020. The period of analysis was from March 1, 2020 to January 10, 2021, and any LTCFs being monitored for exposures or outbreaks were included. If multiple exposures or outbreaks occurred at a facility, the one with the highest attack rate was selected. A total of 74 LTCFs with a COVID-19 exposure or outbreak were included for the descriptive analysis.

For the regression analysis, a total of 48 LTCFs from an initial cohort of 82 LTCFs were included. Appendix B outlines reasons for facility exclusion.

### Data Sources

Various sources were used in our analysis. Data for each exposure or outbreak came from the Fraser Health public health COVID-19 dataset which included information on all COVID-19 cases and exposures/outbreaks in the region. Aggregate case counts for each LTCF exposure or outbreak, the index case, date of the start of monitoring for an exposure or outbreak declaration and date of end of monitoring or date declared over were obtained from this dataset. For calculation of attack rates, LTCF resident and staff census were based on bed capacity and staff payroll information, respectively. To estimate community COVID-19 burden, case data were also used to calculate the incidence rate of COVID-19 in the Local Health Area^23^ of the LTCF for the two weeks prior to the outbreak being declared or the start of monitoring due to an exposure.

Building, organization level and resident characteristics were obtained from the Long-Term Care Facilities Quick Facts Directory from the Office of the Seniors Advocate British Columbia^24^. Lastly, data from the outbreak prevention assessment tool were obtained from the Fraser Health outbreak response teams deployed to the LTCFs following a COVID-19 exposure or outbreak.

### Primary Outcome

The primary outcome variable was outbreak severity measured as the COVID-19 attack rate. The variable consisted of the total number of lab-confirmed COVID-19 cases using the BC Centre for Disease Control case definition^22^ divided by the sum of the resident population (estimated by the number of facility beds) and staff population (staff counts from payroll data from the week of the first exposure). For the descriptive analysis, we analyzed attack rates as a categorical variable (very low, low, medium, high), with cut-points based on the quartile distribution. An additional category of very high was created for LTCFs with an attack rate of >35% based on visual inspection of the data. For the regression analysis, the attack rate was modelled as a continuous variable.

### Predictor variables

We examined various building, organization level and resident population characteristics for the 2019/20 reporting year (Appendix C) as potential risk factors. The complete list of variables examined can be found online in the Long-Term Care Facilities: Quick Facts Directory.^24^ Because this public data source does not include private pay LTCFs and newly opened LTCFs after the 2019/20 reporting year, the data for these predictors used for the regression analysis were obtained from contacting the LTCF for any included LTCF missing data.

From the outbreak prevention assessment performed during the monitoring or outbreak period, we included the score (total number of unmet items) as a predictor variable in the model. We also examined the categories of items in the assessment tool as predictor variables based on whether the facility was in full adherence to all items in a category (yes vs. no).

Other risk factors identified from the literature were also included as predictors. This included community COVID-19 incidence around the LTCF and whether the facility was built prior to 1972, which is when design standards for LTCFs changed in other Canadian jursidictions.^3^ Although for-profit status is considered a strong risk factor in other regions, private pay LTCFs account for a minority of facilities in our region (12/93=13%) and in the regression analysis (3/48=6%), so we did not include for-profit status in our analysis. The Local Health Area^23^ where the LTCF is located was specified as a random effect to account for variation of COVID-19 cases across regions of the health authority. The month of outbreak declaration or start of monitoring was included to account for temporal variation of COVID-19 incidence and adjustments in the protocols for outbreak management over time.

### Statistical Analysis

For the descriptive analysis, each potential risk factor variable was compared across the categorical measure of attack rate (very low, low, medium, high, very high). Given the small sample size and non-normal distribution, the non-parametric statistical tests Kruskal-Wallis test (>2 groups) or Wilcoxon signed-rank test (two groups) for continuous variables and the Fisher’s exact test for categorical variables were utilized. We then modelled the attack rates using negative binomial regression models to account for overdispersion. The LTCF population (total staff and resident beds) were specified as an offset. We created a primary model where the assessment tool score was used as a predictor variable. In secondary models, we assessed the score from each major category of items in the assessment tool as the predictor variable. Model residuals and assumptions were evaluated and met.

## Results

### Descriptive Analysis

During our period of analysis, a total of 145 COVID-19 exposures in 82 LTCFs occurred in the Fraser Health region. Excluding LTCFs where building, organization level and resident population characteristics were not available, the descriptive analysis included 74 facilities (results available in Appendix D). The factors found to be associated with COVID-19 attack rates in LTCFs are summarised in Table 1, along with additional community and outbreak characteristics that were identified as potential risk factors from the literature. Based on the descriptive analysis, the only building characteristics considered as potential risk factors for the regression analysis were the age of the facility and the proportion of single bed rooms. None of the organization level or resident population characteristics were considered for the regression analysis based on the results of the descriptive analysis.

**Table 1.** Descriptive analysis of significant risk factors associated with COVID-19 attack rate

|  | Very Low | Low | Medium | High | Very High | p-value |
| --- | --- | --- | --- | --- | --- | --- |
| N | 19 | 18 | 18 | 12 | 7 |  |
| Community Characteristics |  |  |  |  |  |  |
| Community Incidence within 2 weeks prior to exposure/outbreak (median [IQR]) | 1.22 [0.67, 1.96] | 0.96 [0.52, 2.44] | 1.38 [0.24, 2.24] | 0.33 [0.26, 1.31] | 1.25 [0.80, 2.69] | 0.591 |
| Outbreak Characteristics |  |  |  |  |  |  |
| Index Case Role |  |  |  |  |  |  |
| Resident (%) | 0 (0) | 1 (6) | 7 (39) | 3(25) | 1 (14) | 0.006** |
| Staff (%) | 19 (100) | 17 (94) | 11 (61) | 9 (75) | 6 (86) |  |
| Facility Characteristics |  |  |  |  |  |  |
| % Single Bed Rooms (median(IQR)) | 0.96 [0.93, 1.00] | 0.96 [0.69, 1.00] | 0.97 [0.93, 1.00] | 0.96 [0.91, 0.98] | 0.71 [0.47, 0.88] | 0.048* |
| Facility Opened (median year (IQR)) | 2000 [1991, 2008] | 1989 [1980, 2008] | 2001 [1977, 2008] | 2000 [1990, 2006] | 1974 [1962, 1975] | 0.030* |

\*\*\* p < 0.001; \*\* p < 0.01; \* p < 0.05
Very low: 0%-0.81%, Low: 0.82%-2.9%, Medium: 3.0%-13.2%, High: 13.3%-35%, Very High: >35% -

### Regression Analysis

A total of 48 LTCFs with available data for all potential risk factors were included for the regression analysis. Among these 48 LTCFs, 44 (92%) had a declared outbreak and 4 (8%) were monitored for a COVID-19 exposure with no further transmission. The size of the facilities ranged from a total of 26 beds to 252 beds, with a median of 101 beds. The median attack rate was 2.4%, ranging from 0.2% to 79.2% (IQR: 19.5%). Among residents, the median attack rate was 3.1% (range 0% to 91.3%, IQR:16.6%), and among staff, the median attack rate was 2.2% (range 0% to 83.0%, IQR:7.87%).

From the regression analysis examining the association between potential risk factors and attack rates (Figure 1), the age of the LTCF was significantly associated with increasing attack rates, with LTCFs that opened prior to 1972 having attack rates six times greater than LTCF that opened after 1972 (adjusted rate ratio 5.9 [95% CI 2.3 – 14.9]). The proportion of private rooms in the LTCF, despite being marginally significant in the descriptive analysis, was not associated with attack rates after controlling for other factors.

**Figure 1.**
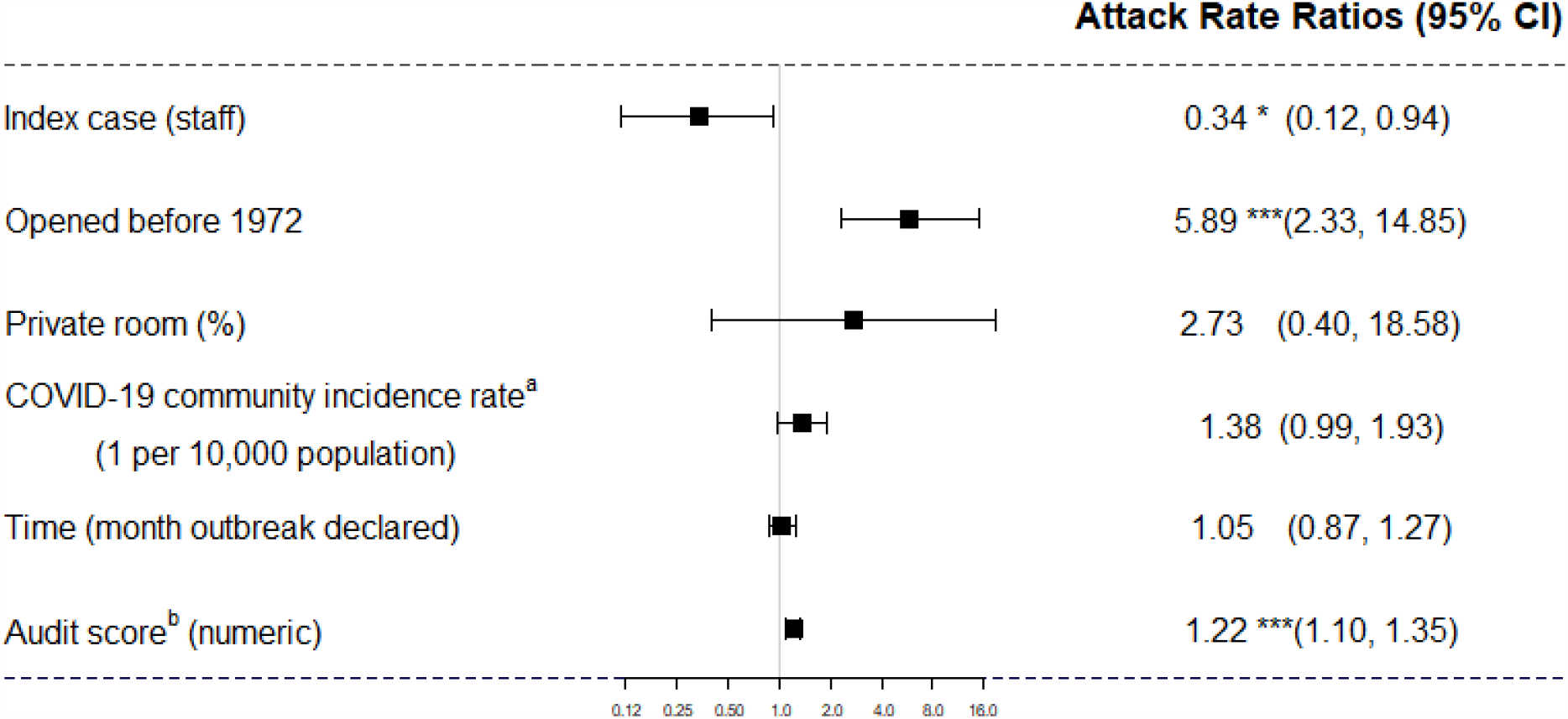
Results of negative binomial regression to examine risk factors associated with COVID-19 attack rates in long-term care facilities in Fraser Health, March 1, 2020 to January 10, 2021 (n=48 unique facilities) Standard errors are heteroskedasticity robust. *** p < 0.001; ** p < 0.01; * p < 0.05 a. Community incidence calculated as incidence rate in the LHA of the LTCF during the two weeks before symptom onset of index case b. Assessment score indicates the number of assessment items that were not met

The assessment tool score, a measure of the IPC and public health measures and readiness of the LTCF for an outbreak, was significantly associated with increasing attack rates. For every item not met in the assessment tool, a 22% increase in the attack rate was observed (adjusted rate ratio 1.2 [95% CI 1.1 – 1.4]). Whether the index case was a staff member or resident was also a significant predictor, with LTCF outbreaks having 66% lower attack rates when the index case identified was a staff member (adjusted rate ratio 0.3 [95% CI 0.1 – 0.9]). The association between COVID-19 community incidence rate and LTCF attack rates was not statistically significant when controlling for other factors, although the model estimate suggested increased attack rates with increasing incidence in the community (adjusted rate ratio 1.4 [95% CI 1.0 – 1.9]).

Secondary models that included a predictor variable for the LTCF’s adherence to various categories of items in the assessment tool (Table 2) demonstrated that if at least one item in the hallway, dining, housekeeping or PPE categories was not met, the LTCFs were more likely to have higher attack rates. In particular, the dining area category had the strongest magnitude of association with attack rates, with an adjusted rate ratio of 3.1 (95% CI 1.4 – 6.8).

**Table 2.** Results of secondary models of negative binomial regression to measure the association between compliance to items in categories of the assessment tool and COVID-19 attack rates in long-term care facilities in Fraser Health, March 1, 2020 to January 10, 2021

| **Category (Not fully compliant with assessment items compared to fully compliant)** | **Attack Rate Ratio<sup>a</sup>** | **95% Confidence Intervals** |
| --- | --- | --- |
| Hallway | 4.03 ** | [1.53, 10.58] |
| Dining | 6.37 *** | [2.70, 15.04] |
| Housekeeping | 4.11 *** | [2.04, 8.31] |
| PPE | 3.10 ** | [1.42, 6.77] |

Standard errors are heteroskedasticity robust. \*\*\* p < 0.001; \*\* p < 0.01; \* p < 0.05
- a. Attack Rate Ratios are adjusted for index case role, facility opened prior 1972, month outbreak declared, community incidence in the Local Health Area, % private rooms. Community specified as random effect.

## Discussion

### Summary of main results

A total of 145 COVID-19 exposures and/or outbreaks occurred from March 1, 2020 to January 10, 2021 in the Fraser Health region. A total of 74 unique LTCF exposures and outbreaks were included in our descriptive analysis which demonstrated that building age, proportion of private rooms, and facility size were associated with increasing outbreak severity as measured by attack rates. Resident population and organization level factors including LTCFs with greater proportion of residents with dementia, wandering, previous complaints/licensing infractions, and nursing care hours were not associated with greater outbreak severity. After controlling for other factors in a regression analysis, building age (LTCFs that opened prior to 1972) and the index case being a resident were associated with increased attack rates at the LTCFs.

As the building age and the index case are non-modifiable, we also examined adherence to IPC and public health measures as modifiable risk factors by including the score from an outbreak prevention assessment tool in our regression model. After controlling for other factors, a linear relationship was found between the number of unmet items in the assessment tool and LTCF attack rates. Specifically, when the LTCF had unmet items in the assessment tool pertaining to measures in the dining area, measures in the hallways, housekeeping/cleaning or PPE availability, there was an increasing likelihood of higher attack rates. These findings support the importance of rigorous adherence to IPC and public health measures but also the complexity of factors that can impact attack rates.

### Explanation of Findings/Comparison to literature

Our findings align with other studies examining factors affecting severity of COVID-19 outbreaks. In terms of LTCF building age, our findings matched those found in Ontario, Canada^3^ – indicating that beds meeting or exceeding “C bed” design standards (Beds that meet or exceed the structural standards of the 1972 Nursing Homes Act Regulation in Ontario) may be a protective factor for COVID-19 transmission. Additionally, the effect of building age may be also due to lack of proper ventilation in older buildings, which has been linked to worsening spread in LTCFs.^25^

Other Canadian studies have found that for-profit status^3,26^ and staffing shortages^26,27^ result in more severe outbreaks in LTCFs. We did not include for-profit status in our analysis due to the small number of private pay LTCFs in our dataset. Staffing shortages were less applicable in our setting, as Fraser Health provided staffing support during an outbreak.

Our findings support a large body of research around the importance of IPC measures in outbreak control and severity.^5,26,27^ Interestingly, the size of the LTCF and the community incidence rate, two factors found to be significantly associated with outbreak severity in other studies^3,26^, were not statistically significant in our analysis, indicating a potential protective effect when IPC and public health measures are in place.

Failure to meet assessment tool items associated with the dining area was a strong risk factor for LTCF attack rates in our jurisdiction. This could be explained by a few reasons: 1) a greater focus on maintenance of IPC measures in patient care areas may have led to poor adherence of IPC measures in dining areas, 2) dining areas may be greater risk areas due to increased bioburden, which combined with poor cleaning/hand hygiene efforts could lead to transmission^28,29^, and 3) as indoor dining areas have been found to be an important setting for transmission in other contexts^30^, dining in the assessment tool may be a surrogate for measures across other parts of the facility that were not directly measured but are globally encompassed within indoor dining practices.

### Future areas for research/action

Our findings support the importance of undertaking a holistic approach to managing COVID-19 outbreaks in LTCFs. In particular, ventilation and building design standards require a more rigorous review as they could inform modified IPC precautions, renovation plans of current LTCFs, and design of future buildings. Some approaches to address ventilation in older LTCFs have been described^25^ and more research is needed on what ventilation counter-measures are most effective. Our analysis did not assess the role of worker policies (e.g., paid sick days) and this is an important area for further assessment.^5^ A Canadian pandemic checklist is in place for LTCF^31,32^ and could be updated to include our findings. The assessment tool used in Fraser Health can also be adapted for other communicable disease outbreaks in LTCFs. We examined exposures and outbreaks prior to or at the start of the COVID-19 vaccination program in British Columbia. Understanding how vaccination efforts reduces outbreak severity will be of interest to understand if the same rigour of outbreak control measures and public health resources will be required in the setting of a highly vaccinated population.

### Strength/Limitations

Our analysis has a number of strengths. First, we used a standardized assessment tool implemented and applied in a uniform fashion across many LTCFs in the region. Second, we were able to include a significant number of LTCF as the Fraser Health region accounted for the majority of the LTCF outbreaks and COVID-19 burden in British Columbia. Third, our final model was able estimate the impact of each risk factor on outbreak severity while controlling for other risk factors. Lastly, we were able to investigate a significant number of building, organization level and resident population characteristics due to a publicly available dataset.^24^

There are, however, limitations to our data and analysis. First, the sample size for our modelling analysis was small, which may have underpowered the analysis and failed to detect significant characteristics. For example, community incidence was found to be a strong determinant for COVID-19 outbreaks in other studies^3,18^ but this was not identified as a major contributor in our model. Second, resident population characteristics were taken from the 2019-2020 cycle and may not accurately reflect characteristics of the population at the time of the outbreak. Third, certain variables that may have contributed to outbreak severity may not have been identified in our analysis as they were not measured in the publicly available data. Fourth, our results may not be generalizable to other jurisdictions where private pay LTCF are more prevalent, as they represent a small proportion of LTCFs in our region. Lastly, due to the rapid and evolving nature of the pandemic response, some outbreak prevention assessments were missing or not carried out during the exposure monitoring or outbreak period. The assessment process underwent continual quality improvement and reconciliation, resulting in a smaller dataset for analysis and potential for introduction of bias in our results.

## Conclusion

Our analysis demonstrates the important modifiable and non-modifiable risk factors associated with COVID-19 outbreaks in LTCFs in our jurisdiction. Our analysis suggests that a higher number of unmet items in a standardized assessment tool deployed routinely at the start of a COVID-19 exposure or outbreak was associated with larger outbreaks. Furthermore, the age of the LTCF and the design standards may be important components to understand outbreak severity and transmission, requiring a standardized evaluation of each LTCF with appropriate interventions.

## Data Availability

The data that support the findings of this study are available on request from the Department of Evaluation and Research Services at Fraser Health. The data are not publicly available due to due to privacy restrictions. However, part of the data that support the findings of this study are available in the Long-Term Care Facilities Quick Facts Directory at https://www.seniorsadvocatebc.ca/long-term-care-quick-facts-directory/

## Authors Contribution

All authors made substantial contributions to the manuscript, including the conception/design (RV, CN, AB), data acquisition (RV, CN, MS), data analysis (RV, CN, MS), and interpretation of data for the work (all authors). RV initially drafted the manuscript, with all authors revising it critically for important intellectual content. All authors provide final approval of the version to be published and agreed to be accountable for all aspects of the work.

## Appendix A. Study Setting

### Outbreak Management Protocol

Fraser Health provides standardized protocols for COVID-19 exposure and outbreak management. When there is a high risk of an outbreak after an exposure at a facility or upon outbreak declaration, an outbreak response lead is deployed to the site. The lead facilitates access to regional expertise and resources, including: 1) Outbreak management expertise from Public Health and the Medical Health Officer; 2) Clinical Nurse Educators for coaching on personal protective equipment (PPE) use and IPC practices ; 3) IPC specialists providing advanced education, assessing IPC measures and providing IPC best practices recommendations; 4) Additional care staff if the LTCF is experiencing staffing shortages; 5) Logistical support for acquiring PPE and supplies; 6) Personnel to facilitate testing; 7) Screeners to ensure active symptom screening of staff, residents, and visitors; 8) and other support as needed.

### Standardized Outbreak Prevention Assessment Tool

The assessment tool was developed by Fraser Health clinical and operational teams with expertise in the LTCF setting in collaboration with IPC and public health, with the purpose of providing a standardized tool for the region that can be used to assess LTCF readiness for COVID-19 outbreaks and to inform outbreak management in the event of an outbreak. The assessment tool includes over 60 public health and IPC measures for prevention and control of COVID-19 outbreaks and pandemic preparedness. The items are grouped into major categories, such as PPE, hand hygiene, etc., and the complete tool can be found in the Supplementary Materials.

The assessment is conducted by Fraser Health staff on site at the onset of an outbreak or a COVID-19 exposure requiring monitoring, and each item in the assessment is marked as met or unmet. An action plan is developed in collaboration with the leadership at the LTCF to address unmet items. An assessment is also done on a routine basis (for outbreak prevention), in the midst of an outbreak (particularly if the outbreak is prolonged), and at the end of an outbreak to ensure that IPC practices and public health measures are maintained even after an outbreak is declared over.

## Appendix B. Exclusion of LTCF for modelling analysis

An initial cohort included 82 LTCFs that experienced an exposure or outbreak from March 1, 2020 to January 10, 2021. Of the 82 facilities, one facility was excluded because the assessment tool data was missing information on the date it was conducted and we could not determine if the assessment was done during the monitoring or outbreak period. An additional 33 facilities were excluded for the following reasons: One facility with an outbreak that occurred prior to the creation of the assessment tool (May 1^st^, 2020); seven facilities with exposures or outbreaks that occurred after revisions to the assessment tool (December 14^th^, 2020) which precluded comparison of data from assessment tools before and after December 14, 2020; and 25 facilities with exposures or outbreaks where an assessment was not conducted or the record was not available in our data system at the time of analysis. If multiple assessments were completed during the monitoring or outbreak period, the assessment with the highest number of unmet items was selected. A total of 48 unique facilities were included in the final dataset.

## Appendix C. Potential risk factors analyzed in descriptive analysis

LTCF building characteristics included age of the facility, the total number of beds, and the number and proportion of private, semi-private, and multi-person rooms. Organization level characteristics included funding type, number of care hours per resident per day, use of contracted services, numbers and rates of licensing complaints and substantiated licensing complaints, and number of infractions found from licensing inspections. Resident population characteristics included demographics, measures of health status such as percent of residents with severe cognitive impairment and average index of social engagement, and utilization of care services such as percent of residents receiving physical therapy.

## Appendix D.

Descriptive analysis of building, organization level, and resident population characteristics and COVID-19 attack rates in long-term care facilities in Fraser Health, March 1, 2020 to January 10, 2021

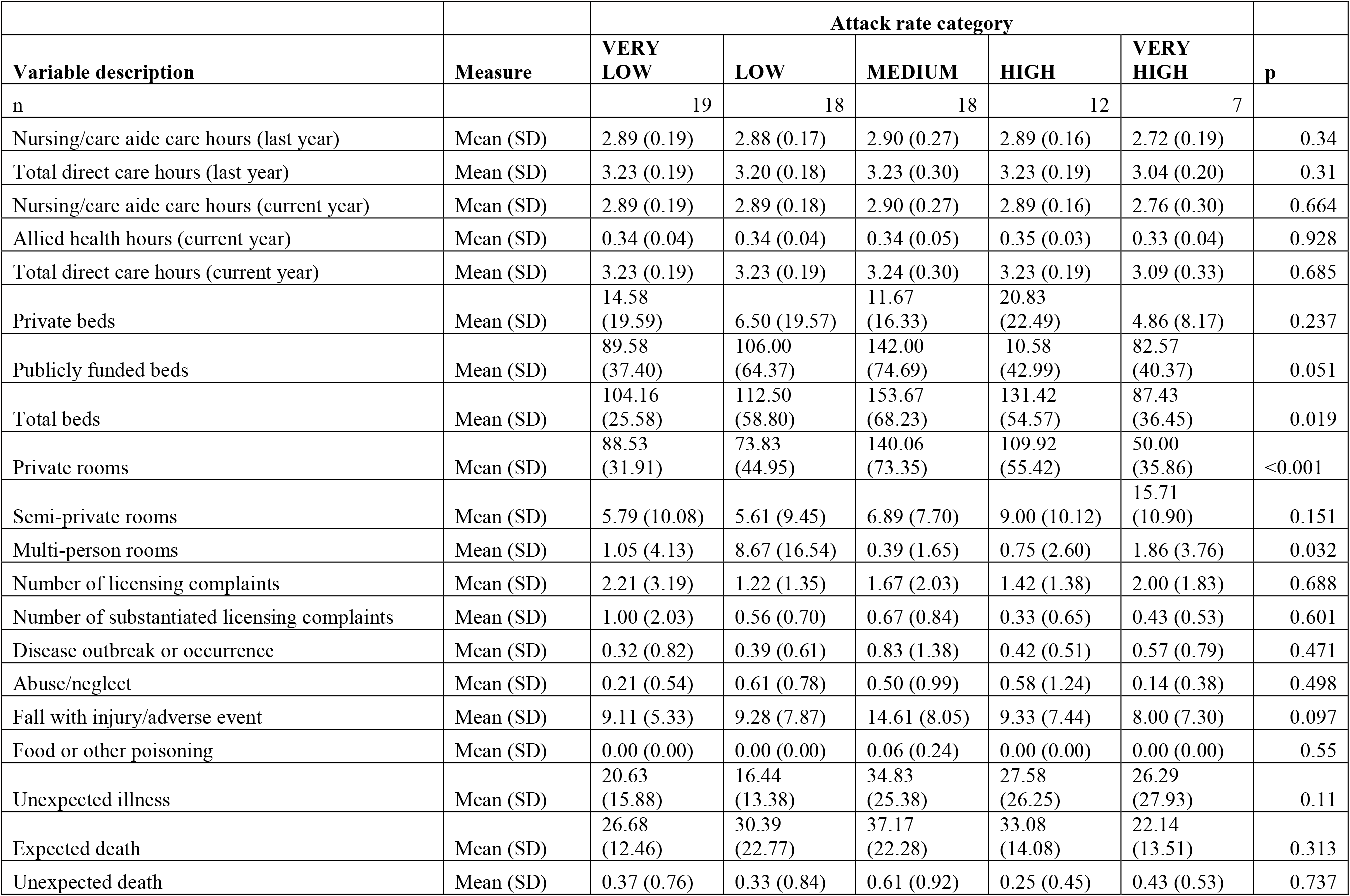
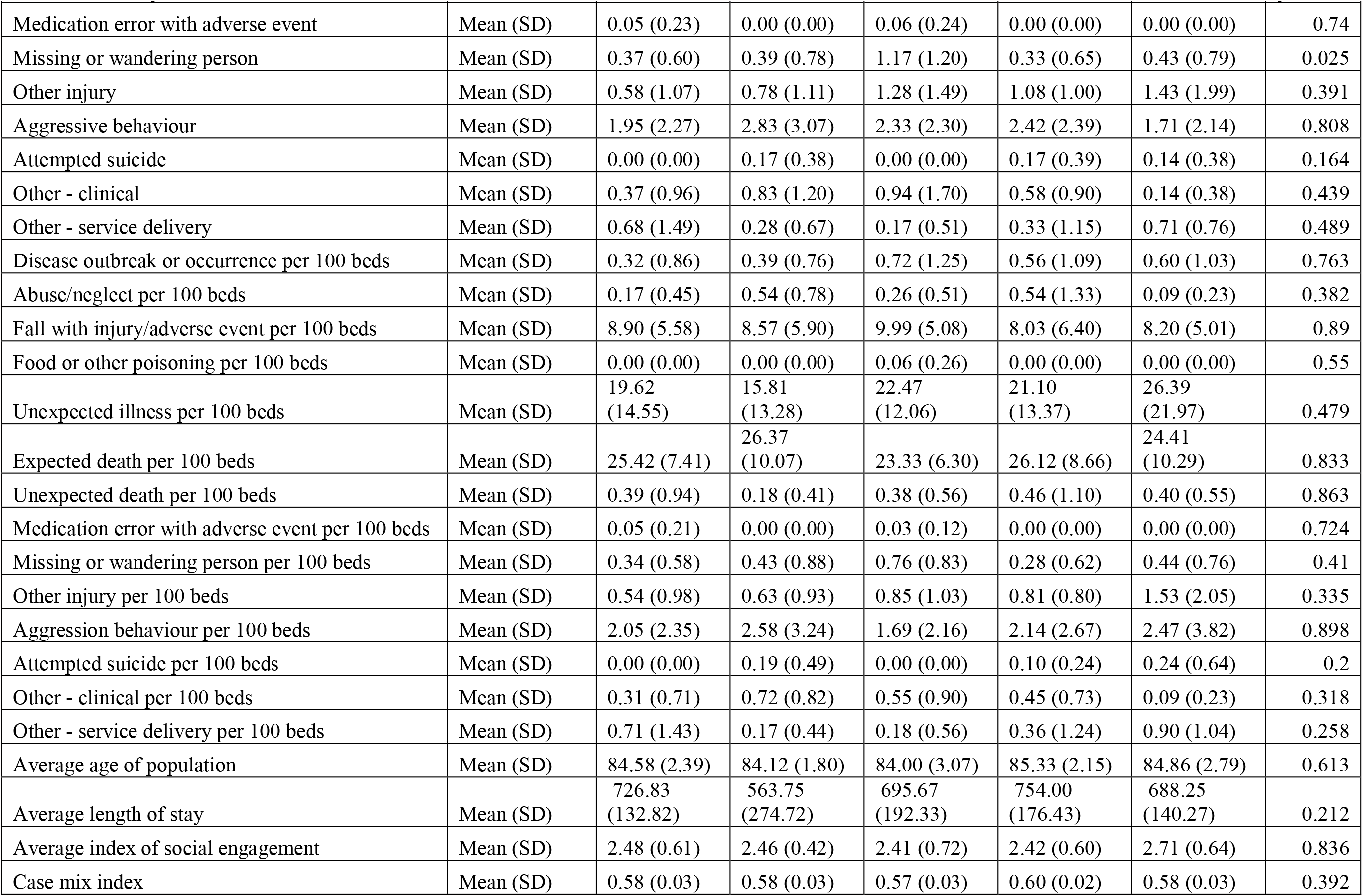
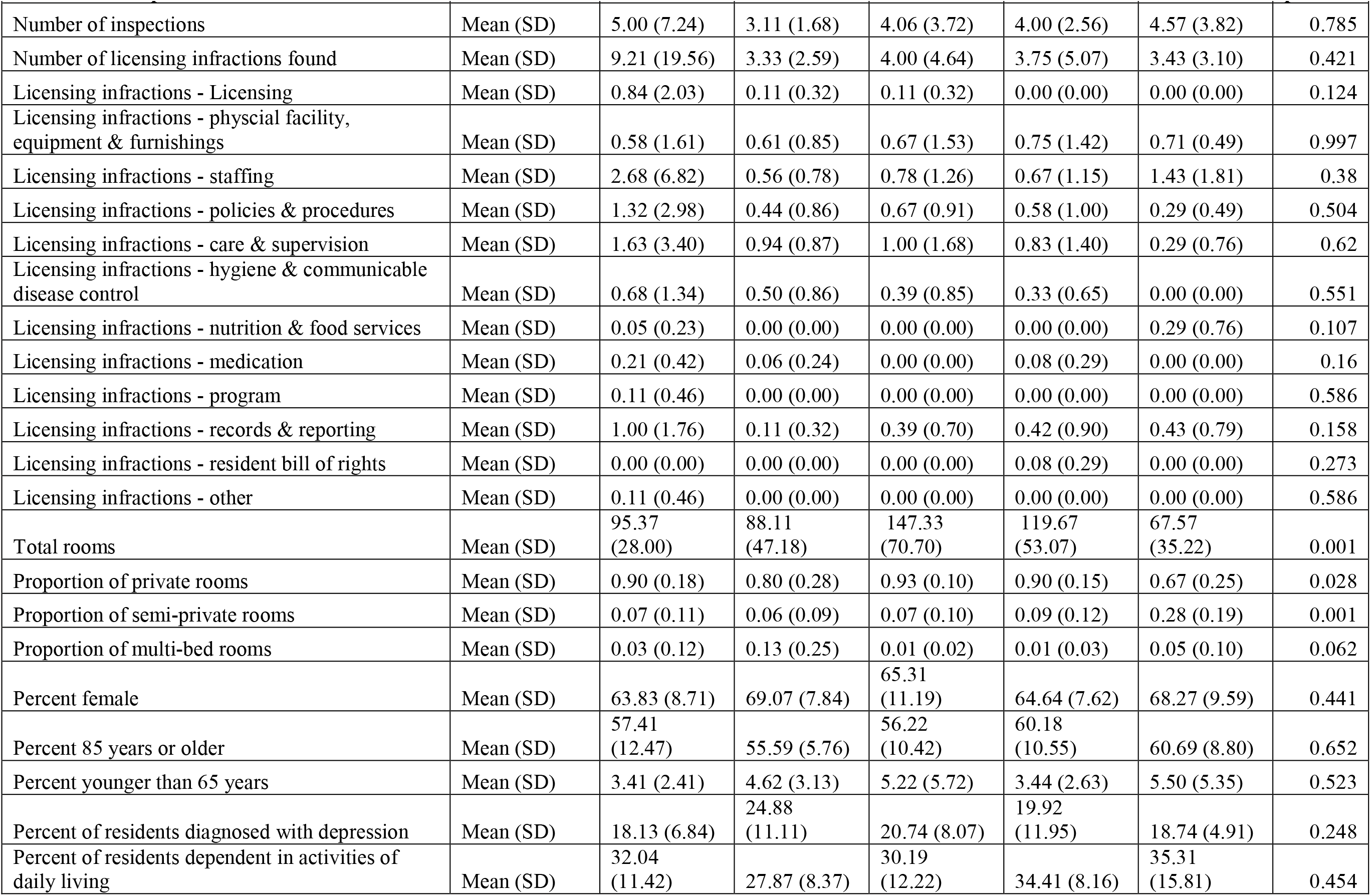
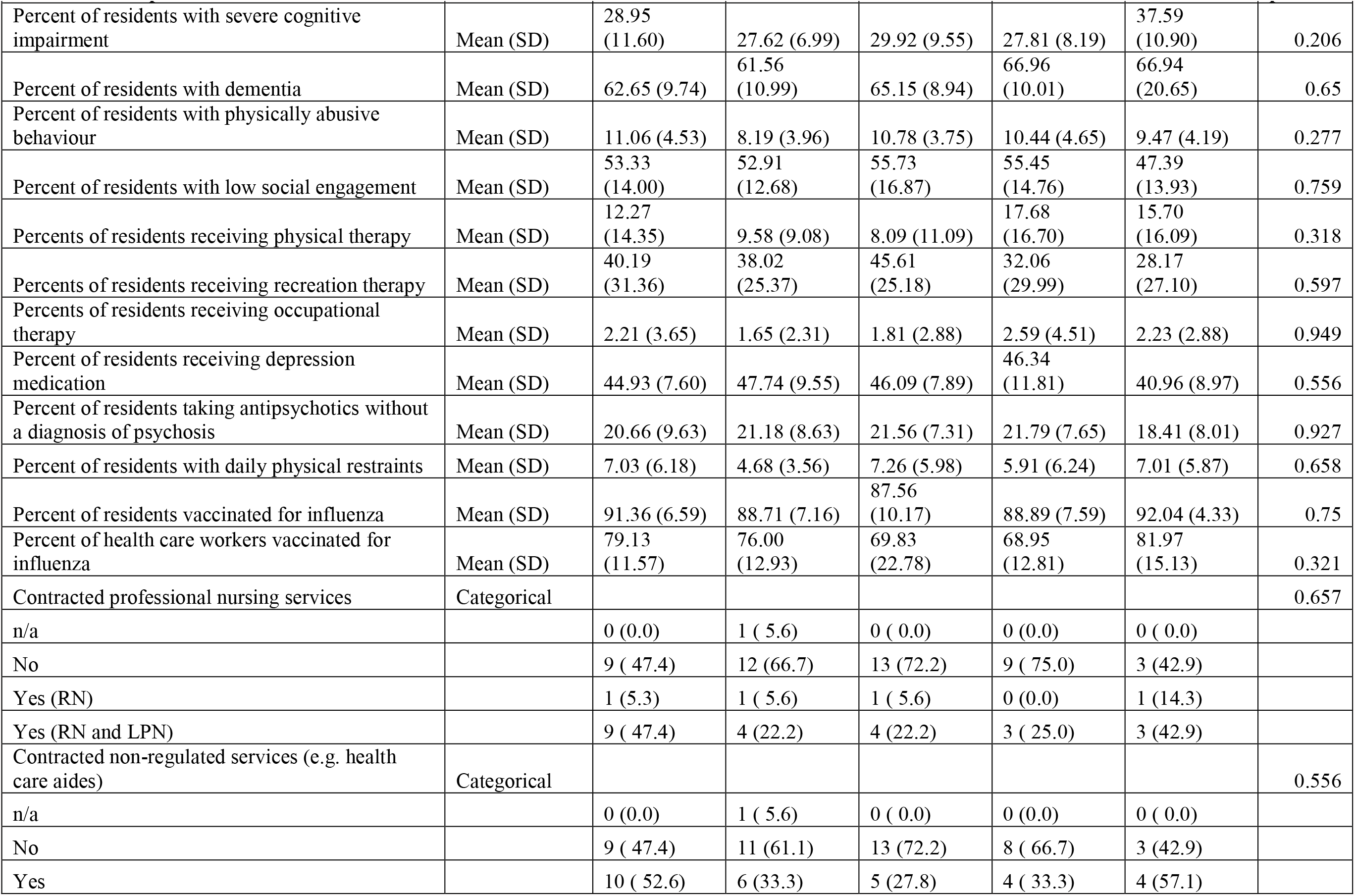
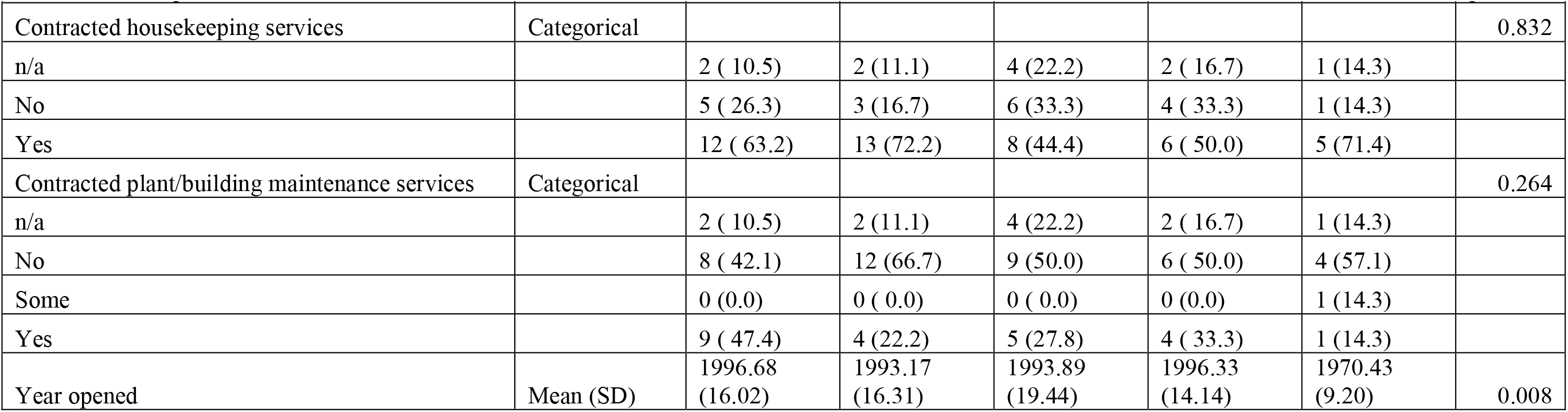

