## Supplementary Materials for "Factors associated with transmission in COVID-19 outbreaks in long-term care facilities"

fraserhealth
Better health. Best in health care.

### COVID-19 Prevention & Outbreak Management Audit Tool For Long-Term Care/Assisted Living/Independent Living - CYCLE #2

| | | |
|---|---|---|
| **Site Name** (if floors have a significantly different population audit all floors): | | **Site Category:** |
| **Site Contact** (e.g. Residential Care Coordinator/Director of Care/Manager): | | **Site Type** |
| ***Auditor Name**: | | **Audit Type:** |
| *If auditor name is not in the list above, please enter it here: | | |
| **Audit Date (mm/dd/yyyy):** | | **Audit Time (hh:mm AM \| PM):** |

**Please Select a "Y" for Yes or "N" for No for each audit question**
**Comments (Optional)**

| # | | Legend:<br>1. ABHR=alcohol based hand rub<br>2. O=Observational/V=Verbal<br>3. High Risk question are identified in Red | QUESTION ANSWER (Y/N) | TIPS for Auditors | Auditor's Comments<br>*(please be as descriptive as possible )* |
|---|---|---|---|---|---|
| **Entrance / Reception / Waiting Area - OBSERVATIONAL** | | | | | |
| 1 | VO | Alcohol-based hand rub is available & accessible & used by all who enter the facility | | Check expiration dates, ensure adequate supply and sites aware of low supply ordering process.Minimum alcohol requirement is 70%. Personal size ABHR in BSTN as wall mounted may not be appropriate | |
| 2 | VO | Screening Process | | Screeners are located at entry /exit points. Active review occuring at entry and exits of all staff and visitors. | |
| 3 | VO | Signage- at entrance & exit( s) , precautions such as physical distancing, PPE, refrain from entering if ill | | There is signage advising staff and visitors (including BC Emergency Services, couriers), single entrance, IPC precautions eg: respiratory etiquette, physical distancing, PPE for all staff, clients, visitors. Posters are available and visible and instructing staff and visitors to restrain from entering if ill. | |
| 4 | VO | Single control access point is recommended for screening and access to building * sites may have a variety of stand alone buildings so there will be more than one staff access point e.g.: campus of care. | | Please ensure compliance with signage, screening and physical distancing. CAUTION and pay attention with shared corridors and connected buildings that maintaining physical distancing is possible. Sites may have more than one entrance point such as one for staff only and one for visitors. Signage must be in place if more than one entrance point. The main principle is that there is a signage that is visible at all entrance points. | |
| 5 | VO | Ensure PPE availability eg: masks & protective eye wear are available at the entrance. | | Visible and supply is adequate. Ensure staff are donning their mask & eye wear & safe maintenance of physical distancing | |
| **Visits - Essential** | | | | | |
| 6 | VO | 1 essential visitor **during outbreak** (e.g. compassionate visits only , actively dying), visitors must be asymptomatic. | | Essential visits only when in outbreak. RESOURCE: Essential Visitor protocol | |
| **Visits -Family - Social** | | | | | |
| 7 | V O | Family/social visits require a written plan and screening process, vistors are wearing a mask ( **can be personal cloth or surgical procedural mask*** ) , must be asymptomatic | | Social/family visits poster visible, identifiable designated visiting areas, written plan ( 1 designated visitor) for scheduling and registering visitors, ensure process of review of PPE and IPC standards has been completed. **OUTBREAK : CLOSE FAMILY OR SOCIAL VISITS and comment** | |
| **Personal Care Services (eg: Hairdressing)** | | | | | |

fraserhealth
Better health. Best in health care.

### COVID-19 Prevention & Outbreak Management Audit Tool For Long-Term Care/Assisted Living/Independent Living - CYCLE #2

| | | |
|---|---|---|
| **Site Name** (if floors have a significantly different population audit all floors): | | **Site Category:** |
| **Site Contact** (e.g. Residential Care Coordinator/Director of Care/Manager): | | **Site Type** |
| ***Auditor Name**: | | **Audit Type:** |
| *If auditor name is not in the list above, please enter it here: | | |
| **Audit Date (mm/dd/yyyy):** | | **Audit Time (hh:mm AM \| PM):** |

**Please Select a "Y" for Yes or "N" for No for each audit question**
**Comments (Optional)**

| # | | **Legend:**<br>**1. ABHR=alcohol based hand rub**<br>**2. O=Observational/V=Verbal**<br>**3. High Risk question are identified in Red** | **QUESTION ANSWER (Y/N)** | **TIPS for Auditors** | **Auditor's Comments**<br>*(please be as descriptive as possible )* |
|---|---|---|---|---|---|
| 8 | VO | Hairdressing & Personal Care Services ( written plan and site specific) | | Sites may permit one hairdresser to provide hairdressing services as long as a personal care service has provided a written plan to facility,ensure process of screening, review of PPE and IPC standards has been completed, registration and monitoring of services. Worksafe BC protocols must align https://www.worksafebc.com/en/about-us/covid-19updates/covid-19-returning-safe-operation/personal-services. See toolkit for other resources | |
| 9 | VO | Furniture in common areas is clean ( wipeable furniture should be cleaned twice a day( all high touch areas) . Fabric furniture should be clean, not torn and stained and cleaned q3 months | | Furniture is is not torn, broken or heavily soiled. If so it must be repaired, replaced or cleaned. Reference: Environmental cleaning guidelines & best practices by FH IPC & BC CDC & PIC NET ( see p. 10-11 ) **https://www.picnet.ca/wp-content/uploads/British-Columbia-Best-Practices-for-Environmental-Cleaning-for-Prevention-and-Control-of-Infections-in-All-Healthcare-Settings-and-Programs.pdf** * note in AL and IL environments this may be difficult when promoting a home like environment and not all furniture is wipeable eg: non fabric particularly in common areas. if that is the case ensure the furniture is clean and is reguarly cleaned - minmum of Q3 months. | |
| **Staff Screening** | | | | | |
| 10 | VO | Active Screening 2 x per shift: Beginning and during shift for all staff. Screen staff for: symptoms (i.e. fever, new or worsening cough, new or worsening shortness of breath, sore throat, and nausea /vomiting and diarrhea); travel outside of Canada, and/or; contact with confirmed COVID-19 case. | | ACTIVE SCREENING of all staff: follow BC CDC guidelines for screening at beginning of shift and during shift . Staff screening of each other must occur and it must be documented during their shift. **FH Screeners can be deployed in an outbreak situation and screening will occur 2 x shift beginning and during shift.** | Staff prefill their screening forms prior to reporting to work and present them to the screener who checks their temperature. The staff member then takes the form to their unit for the mid shift screen. |
| 11 | VO | All staff have been provided with information on how to self-monitor for symptoms. | | Poster for staff monitoring in locations that are visible to staff ( eg: entrance to building, staff lounge) , staff have been educated on self monitoring and it is reviewed regularly by site leadership with their teams | |
| 12 | VO | Documentation of findings of self report and checklists are kept in a binder or folder by site leaders. | | | |

### COVID-19 Prevention & Outbreak Management Audit Tool For Long-Term Care/Assisted Living/Independent Living - CYCLE #2

| | | |
|---|---|---|
| **Site Name** (if floors have a significantly different population audit all floors): | | **Site Category:** |
| **Site Contact** (e.g. Residential Care Coordinator/Director of Care/Manager): | | **Site Type** |
| ***Auditor Name**: | | **Audit Type:** |
| *If auditor name is not in the list above, please enter it here: | | |
| **Audit Date (mm/dd/yyyy):** | | **Audit Time (hh:mm AM \| PM):** |

**Please Select a "Y" for Yes or "N" for No for each audit question**
**Comments (Optional)**

| # | | Legend: 1. ABHR=alcohol based hand rub 2. O=Observational/V=Verbal 3. High Risk question are identified in Red | QUESTION ANSWER (Y/N) | TIPS for Auditors | Auditor's Comments (*please be as descriptive as possible*) |
|---|---|---|---|---|---|
| 13 | VO | If staff are symptomatic, they are aware of process, to report to the supervisor immediately, remove themselves from work, refer to testing assessment center and self-isolate pending results. | | Ensure any staff member with even mild symptoms has access to a supervisor who can quickly reassign the work and release the employee to go home. Check there are posters for staff monitoring in locations that are visible to staff ( eg: entrance to building, staff lounge). | |
| **Resident/Client/Tenant Screening & Swabbing** | | | | | |
| 14 | VO | Active Screening' twice a day (2 x a day) as per BC CDC guidelines of all residents/persons for: symptoms (i.e. fever, new or worsening cough, new or worsening shortness of breath, sore throat, nausea /vomiting and diarrhea ); travel outside of Canada, and/or; contact with confirmed COVID-19 case. | | Nausea/vomiting and diarrhea are symptoms recognized as per MHO guidance and FH screening tool, BC CDC guidelines. | |
| 15 | VO | Is there an awareness of what to do if a resident/client/tenant becomes symptomatic, what is the process for isolating , accessing equipment and instruction for COVID-19 swabbing and notifying Public Health. | | Signage for droplet precautions is available, process for dedicated equipment eg: PPE available, prepared kits, and carts are available for placing supplies. If there is a delay can contribute to transmisson risk | |
| 16 | VO | Are site leadership aware of how to obtain nasopharyngeal ( NP) swabs and as appropriate to do NP swab & package and transport swab. Please see BCCDC guidelines and COVID toolkit. | | Refer to BCCDC website. Home testing for AL IL, private sites will receive training on how to obtain NP swabs ( back up is home testing until trained) . See KYI for TDG- staff must be certified in how to package the swab. | |
| **Resident/Client/Tenant Movement** | | | | | |
| 17 | V | Ensure awareness of transfer, admission, readmission requirements | | Please review & ensure availability of Fraser Health COVID-19 Screening Process/Algorithm, and admission & transfer algorithm for acute and community | |
| 18 | O/V | Have all group activities into the community or from the community into the site stopped for LTC | | This includes that no community groups may enter. Day programs co-located with LTC sites are temporarily closed. AL/IL tenants and clients may enter community as considered under general public health measures. | |
| 19 | V | Residents of LTC go into the community only for essential medical needs only . Tenants of AL & IL can engage in social and external activities aligned with general public health guidance. This may include non essential outings such obtaining essential groceries. | | Please see if sites have reviewed and are aware of the KYI and algorithm for transfer for essential medical visits for LTC& AL . | |
| 20 | | Resident/client/tenant may be permitted to receive essential clinical services with a onsite written plan of process | | Please see if sites have reviewed and are aware of the KYI for essential clinical services ( e.g.: foot care or dental) | |
| **Recreation** | | | | | |

fraserhealth
Better health. Best in health care.

### COVID-19 Prevention & Outbreak Management Audit Tool For Long-Term Care/Assisted Living/Independent Living - CYCLE #2

| | | |
|---|---|---|
| **Site Name** (if floors have a significantly different population audit all floors): | | **Site Category:** |
| **Site Contact** (e.g. Residential Care Coordinator/Director of Care/Manager): | | **Site Type** |
| ***Auditor Name**: | | **Audit Type:** |
| *If auditor name is not in the list above, please enter it here: | | |
| **Audit Date (mm/dd/yyyy):** | | **Audit Time (hh:mm AM \| PM):** |

**Please Select a "Y" for Yes or "N" for No for each audit question**
**Comments (Optional)**

| # | | **Legend:**<br>**1. ABHR=alcohol based hand rub**<br>**2. O=Observational/V=Verbal**<br>**3. High Risk question are identified in Red** | **QUESTION ANSWER (Y/N)** | **TIPS for Auditors** | **Auditor's Comments**<br>(*please be as descriptive as possible* ) |
|---|---|---|---|---|---|
| 21 | O/V | Congregate activities such as recreation are occurring with 2 meter physical distancing and 'no touch' practices being maintained | | Recreation can be occurring as long as 1-1 physical distancing of 2 meters in place and staff are wearing surgical/procedural mask & protective eye wear. Maximize physical separation as much as possible within the confines of the physical environment. E.g. increase physical separation while dining, stagger meal times, /meal trays as appropriate. * there are supporting documents related to supporting residents with dementia who may be restless or having trouble adhering to expectations. | |
| **Resident/Tenant/Client Information** | | | | | |
| 22 | V | All resident/persons contacts are up-to-date, including family and medical practitioners. | | | |
| 23 | V | Does the leadership have a mechanism of contacting family with sharing of site updates or other information. | | | |
| 24 | V | All resident/persons goals of care are documented in the advance care plan and the Medical Orders Scope of Treatment ( MOST) are up-to-date (recent updating with representative/substitute decision maker). | | In AL MOST is not a requirement but best practice is to encourage goals of care conversations and document in advance care planning record. May be completed by AL Community Care professional. If not applicable please leave blank and add note in comments | |
| **Staffing** | | | | | |
| 25 | V | Is there a written staffing plan of how to obtain staff in the event of a critical shortage. | | this is an ask for do they or don't they have a staffing plan. | |
| 26 | V | Is there evidence that all staff have had ongoing retraining (q 3 months) on infection control ( this includes hand hygiene, PPE, and additional precautions). | | Sites must demonstrate by showing sign in sheets for education. Additoinal precautions ( airborne, contact, & droplet) | |
| **Nursing Stations/ Algove/Office ( can be on multiple floors and all need to be checked) - OBSERVATIONAL** | | | | | |
| 27 | O | Clean, orderly and free of clutter. | | Non clutter to enable housekeeping to clean horizontal surfaces. | |
| 28 | O | There is no food and drink. | | No open food or drink, no potlucks, OUTBREAK: Staff may not bring in personal water bottles | |
| 29 | O | Clear of personal items. | | Hand bags, lunch kits, jackets, sweaters. | |
| 30 | O | Dedicated Hand hygiene sink on unit or floor or Alcohol Based Hand Rub (ABHR) is available & accessible (e.g. container is not empty). | | Check for expiration dates, low supply ordering process. | |
| **Soiled Utility Room/Housekeeping Room/Laundry Room** | | | | | |

### COVID-19 Prevention & Outbreak Management Audit Tool For Long-Term Care/Assisted Living/Independent Living - CYCLE #2

| | | |
|---|---|---|
| **Site Name** (if floors have a significantly different population audit all floors): | | **Site Category:** |
| **Site Contact** (e.g. Residential Care Coordinator/Director of Care/Manager): | | **Site Type** |
| ***Auditor Name**: | | **Audit Type:** |
| *If auditor name is not in the list above, please enter it here: | | |
| **Audit Date (mm/dd/yyyy):** | | **Audit Time (hh:mm AM \| PM):** |

**Please Select a "Y" for Yes or "N" for No for each audit question**
**Comments (Optional)**

| # | | **Legend:**<br>**1. ABHR=alcohol based hand rub**<br>**2. O=Observational/V=Verbal**<br>**3. High Risk question are identified in Red** | **QUESTION ANSWER (Y/N)** | **TIPS for Auditors** | **Auditor's Comments**<br>*(please be as descriptive as possible )* |
|---|---|---|---|---|---|
| 31 | O | Door to room is kept closed and restricted access. | | | |
| 32 | O | Hand hygiene sinks have paper towel and plain liquid soap dispensers in close proximity (e.g. ensure container is not empty). | | Hands free is optimal and best practice but not mandatory. | |
| 33 | O | Use of precautions when doing Laundry and observations in Laundry room as a common area (tips apply particularly in IL as a campus of care). | | Contaminated laundry should be placed into a laundry bag or basket with a plastic liner and should not be shaken. Gloves and a surgical or procedural mask, or if not available, a non-medical mask ( cloth mask), should be worn when in direct contact with contaminated laundry. Clothing and linens belonging to the ill person can be washed together with other laundry, using regular laundry soap and **hot water (60-90°C)**. Laundry should be thoroughly dried. Hand hygiene should be performed after handling contaminated laundry and after removing gloves. If the laundry container comes in contact with contaminated laundry, it can be disinfected using the diluted bleach solution. | |
| 34 | O | The hand hygiene sink is completely free from surrounding clutter. | | | |
| **Clean and Sterile Supply Room** | | | | | |
| 35 | V | Sterile storage has a designated area and is limited to only staff access and door to room is kept closed. | | | |
| 36 | V& O | PPE are kept in secured area. | | Supplies are adequate in the event of an outbreak (minimum of 3 day supply) which includes surgical/procedural masks, protective eye wear, gloves and gowns. Are kits/carts * not cardboard boxes- prepared in advance is reccomended if not ready to go. Will be covered in flu school. | |
| **Hallway - Observational** | | | | | |
| 37 | O | ABHR is available and accessible outside residents/tenants room or common area in AL | | ABHR = alcohol based hand rub ; please check for expiration date. One per room available on a table due to wall mount shortage is acceptable | |
| 38 | O | Free of clutter- carts, wheelchairs, equipment. | | There is a clear separation between clean and dirty equipment/items. Note: some sites are short on wall mounted ABHR and may have it placed on a table or cart in hallway outside rooms. | |
| **Dining areas e.g. kitchenettes, servery, eating area** | | | | | |

### COVID-19 Prevention & Outbreak Management Audit Tool For Long-Term Care/Assisted Living/Independent Living - CYCLE #2

| | | |
|---|---|---|
| **Site Name** (if floors have a significantly different population audit all floors): | | **Site Category:** |
| **Site Contact** (e.g. Residential Care Coordinator/Director of Care/Manager): | | **Site Type** |
| ***Auditor Name**: | | **Audit Type:** |
| *If auditor name is not in the list above, please enter it here: | | |
| **Audit Date (mm/dd/yyyy)**: | | **Audit Time (hh:mm AM \| PM):** |

**Please Select a "Y" for Yes or "N" for No for each audit question**
**Comments (Optional)**

| # | **Legend:**<br>**1. ABHR=alcohol based hand rub**<br>**2. O=Observational/V=Verbal**<br>**3. High Risk question are identified in Red** | | **QUESTION ANSWER (Y/N)** | **TIPS for Auditors** | **Auditor's Comments**<br>*(please be as descriptive as possible )* |
|---|---|---|---|---|---|
| 39 | O | Kitchenettes are closed. | | **Food/beverage service must be monitored , scheduled and provided by AL staff .** NO SELF SERVICE IS ALLOWED. NOT PERMITED IN OUTBREAK | |
| 40 | O | Ensure scheduled cleaning of all surfaces in dining area after every meal, following every sitting, minimum of twice a day. | | Sites need to demonstrate the schedule for cleaning. **In outbreak Dining rooms must be closed**. NO PLEXI GLASS. | |
| 41 | O | Environmental controls are in place to ensure physical distancing (2 metre distancing). | | congregate dining is a high risk activity and must have an environment large enough to maintain 2 meter physical distanc. Sites may schedule/stagger rotating dining times to accomodate increasing physical separation while dining, use individual meal trays as appropriate. *refer sites as needed to documents supporting residents with dementia who may be restless or having trouble adhering to expectations. | |
| 42 | O | Hand Hygiene sink/ABHR is available (e.g. container is not empty). | | Please check all areas even if closed. Ask to see the area eg: closed dining room. | |
| 43 | V | There is a process for cleaning residents/tenants hands before/after meal time. | | Ensure wipes ( ABHR type) eg: Sani /Wet ones are used as needed in LTC and assistance is provided or ABHR is available in both LTC and AL. | |
| 44 | O | There is signage indicating step-by-step guide to proper hand hygiene (HH). | | Poster reminders " steps to perform HH" . | |
| 45 | O/V | Furniture cleanable and able to withstand disinfection with hospital grade disinfectant. | | **See element # 9 for details** . | |
| **Housekeeping** | | | | | |
| 46 | O/V | Is enhanced cleaning in place for the entire facility, with high touch surfaces cleaned and disinfected twice a day (6-8 hours between cleaning). | | For example high touch surfaces are counters, door knobs, faucets, furniture arms, back and seat if wipeable. UVG Marker compliance is a monthly check of **90%** . Enhanced cleaning standards are set by regular audits (minimum of monthly in prevention; weekly if outbreak; increase to 3 x week if less than 90% on audit score in prevention ) * owned and operated may be doing regular audits weekly. | |

fraserhealth
Better health. Best in health care.

### COVID-19 Prevention & Outbreak Management Audit Tool For Long-Term Care/Assisted Living/Independent Living - CYCLE #2

| | | |
|---|---|---|
| **Site Name** (if floors have a significantly different population audit all floors): | | **Site Category:** |
| **Site Contact** (e.g. Residential Care Coordinator/Director of Care/Manager): | | **Site Type** |
| ***Auditor Name**: | | **Audit Type:** |
| *If auditor name is not in the list above, please enter it here: | | |
| **Audit Date (mm/dd/yyyy)**: | | **Audit Time (hh:mm AM \| PM):** |

**Please Select a "Y" for Yes or "N" for No for each audit question**
**Comments (Optional)**

| # | | **Legend:**<br>**1. ABHR=alcohol based hand rub**<br>**2. O=Observational/V=Verbal**<br>**3. High Risk question are identified in Red** | **QUESTION ANSWER (Y/N)** | **TIPS for Auditors** | **Auditor's Comments**<br>*(please be as descriptive as possible )* |
|---|---|---|---|---|---|
| 47 | O/V | Regularly scheduled cleaning of housekeeping equipment e.g.: mop heads; toilet brushes, according to protocol. | | Best practice is that there is a dedicated toilet brushes per resident room as appropriate ( check routine practices); mop heads are laundered daily & dried thoroughly before storage. **CHECK** BCCDC website **ENVIRONMENTAL STANDARDS VIA PICNET.** | |
| 48 | | Garbage bins are recommended to be handsfree in all areas | | Garbage bins are considered dirty . Future purchases are suggestive of hands free | |
| 49 | | Staff rooms: Cleaning between use, after use, between shifts, ensuring environmental controls for physical distancing , scheduled and staggered breaks. | | Available ABHR; enhanced cleaning guidelines to be enforced due to high transmission risk ( cleanser available for staff to clean) ; facilities can remove chairs /space accordingly to accommodate physical distancing requirements. Microwaves ( interior /exterior clean daily ) and fridges exterior clean daily, interior clean weekly | |
| 50 | O | Sites demonstrate and show what they are using for cleaning -should be .5% accelerated hydrogen peroxide or hospital grade disinfectant eg: bleach wipes/cavi wipes. Health Canada approved disinfectants for COVID-19. | | Sites must show you what they are using for cleaning/refer them to Health Canada website. The disinfectant does not have to be in the form of a wipe. See BCCDC specifications and link below<br>• Disinfectants should be classed as a hospital grade disinfectant and registered in Canada with a Drug Identification Number (DIN)<br>• Follow product instructions for dilution, wet contact time and safe use (e.g., use of PPE and proper ventilation).http://www.bccdc.ca/Health-Info-Site/Documents/Environmental_Service_Providers_Health_Care.pdf | |
| **Reprocessing -Cleaning instructions** | | | | | |

fraserhealth
Better health. Best in health care.

### COVID-19 Prevention & Outbreak Management Audit Tool For Long-Term Care/Assisted Living/Independent Living - CYCLE #2

| | | |
|---|---|---|
| **Site Name** (if floors have a significantly different population audit all floors): | | **Site Category:** |
| **Site Contact** (e.g. Residential Care Coordinator/Director of Care/Manager): | | **Site Type** |
| ***Auditor Name**: | | **Audit Type:** |
| *If auditor name is not in the list above, please enter it here: | | |
| **Audit Date (mm/dd/yyyy):** | | **Audit Time (hh:mm AM \| PM):** |

**Please Select a "Y" for Yes or "N" for No for each audit question**
**Comments (Optional)**

| # | | **Legend:**<br>**1. ABHR=alcohol based hand rub**<br>**2. O=Observational/V=Verbal**<br>**3. High Risk question are identified in Red** | **QUESTION ANSWER (Y/N)** | **TIPS for Auditors** | **Auditor's Comments**<br>*(please be as descriptive as possible )* |
|---|---|---|---|---|---|
| 51 | O/V | Cleaning instructions ( work) are available for all shared equipment and recreational equipment (e.g. lifts, walkers, wheelchairs, slings both sit to stand and ceiling lifts, shower chair, tub; recreation tools such as bowling equipment, instruments) | | Ensure work/cleaning instructions are visible and are being cleaned appropriately. Sit to stand slings **cannot** be used for more than one resident if soiled-they must be laundered between use. They cannot just be wiped off and sprayed with disinfectant if fabric sections-must be laundered. If wipeable no fabric sections, CAVI WIPES are suitable. Encourage disposable slings particularly if in outbreak. * Action planning may entail purchasing more slings. Ceiling lifts are resident specific. There is a policy and procedure ( **link to policy and procedure**) for cleaning and<br>disinfection of environmental surfaces and<br>shared equipment (e.g., commodes, wheelchairs,) | |
| 52 | O/V | There is no sharing of equipment , wash basins or supplies between residents/persons without appropriate cleaning and disinfection between use. | | Basins must be dedicated. No shared use prior to cleaning and disinfected and clear labelling is required. **If disposable equipment is used, it is single-use** | |
| **PPE** | | | | | |
| 53 | V | PPE is available, stored appropriately and accessible for all staff. Staff are also provided what is required for direct care to residents. | | Surgical procedural mask, protective eye wear, (gown and gloves for droplet precautions) and must be placed on proper PPE station /cart with a wipeable surface that is reusable. PPE should not be placed on a chair or open cardboard box with no lid. | |
| 54 | O/V | Is a three day supply of PPE available on site (including surgical/procedural masks, gloves, gowns, and eye protection)? | | Are they aware of how to order and have activated an order if necessary /plan to replenish supplies, **cloth masks are not permitted for staff.** | |
| 55 | O | Staff are wearing recommended PPE when in resident common areas & when providing direct care. The recommended PPE must be worn at all times in these circumstances. They are surgical/procedural mask and protective eye wear. | | **Cloth masks are not permitted for staff.** | |
| 56 | O | Resident/tenant/client who is on droplet precautions staff must be in PPE ( includes mask, eye wear, gowns and gloves). | | Ensure staff are following FH recommendations when to discard used PPE and that there is a dedicated cart with appropriate PPE dedicated. | |
| 57 | V/O | All staff must have been trained in donning and doffing procedures-confirm there is a documented process of PPE donning and doffing auditing and ask to see most recent audit. Ensure training has occurred q3 months | | Auditor can ask for staff retraining sign in sheets , ensure they are available and site audits are in place. Gown and gloves are not to be worn outside of resident environment unless worn for completion of task? **(eg: taking soiled laundry to the dirty utility room)** | |

fraserhealth
Better health. Best in health care.

### COVID-19 Prevention & Outbreak Management Audit Tool For Long-Term Care/Assisted Living/Independent Living - CYCLE #2

| | | |
|---|---|---|
| **Site Name** (if floors have a significantly different population audit all floors): | | **Site Category:** |
| **Site Contact** (e.g. Residential Care Coordinator/Director of Care/Manager): | | **Site Type** |
| ***Auditor Name**: | | **Audit Type:** |
| *If auditor name is not in the list above, please enter it here: | | |
| **Audit Date (mm/dd/yyyy)**: | | **Audit Time (hh:mm AM \| PM):** |

**Please Select a "Y" for Yes or "N" for No for each audit question**
**Comments (Optional)**

| # | | **Legend:**<br>**1. ABHR=alcohol based hand rub**<br>**2. O=Observational/V=Verbal**<br>**3. High Risk question are identified in Red** | **QUESTION ANSWER (Y/N)** | **TIPS for Auditors** | **Auditor's Comments**<br>*(please be as descriptive as possible )* |
|---|---|---|---|---|---|
| 58 | OV | Review Infection Prevention Standards ( IPC) standards for isolation ( droplet precautions) in the event a resident/tenant became symptomatic, including swabbing for COVID-19 and notifiying Public Health. Ensure observed signage outside ( at door) / in multi bedrooms ensure signage is posted above the residents bed. Ensure PPE carts are available and placed outside room for any resident or tenant currently on droplet precautions. | | Please ask do you have any residents who are on droplet precautions and if yes please go and visit where resident room is and if signage is posted and PPE is accessible and available. **PPE SIGNAGE IS ACCREDITATION CANADA STANDARD ( * AL sites are not accredited)** | |
| **Hand Hygiene ( HH)** | | | | | |
| 59 | V/O | "Your 4 Moments of Hand Hygiene poster" is visible in all resident /tenant care areas | | The 4 moments HH practice before resident/environment contact; before aseptic procedure; after blood and body fluid exposure risk; after resident/environment/client/tenant contact. Observe hand hygiene practices. **In AL placing of posters must be on the wall outside of the suites.** | |
| 60 | V/O | Hand Hygiene (HH) audits are completed at a minimum monthly (NOTE: **Hand hygiene audit frequency will increase to three times a week during outbreaks or daily if a complex outbreak.**) . Observe staff hand hygiene practices | | Sites must show documentation and auditor observe what their compliance results are and they need to be 80% or higher. **If sites are under 80% increase audit frequency to weekly until compliant.** AL/IL sites may not meet this requirement however they must ensure they are aware of audits and importance and may see that sites are using a non FH tool. Non FH tool must include 4 moments HH | |
| 61 | V/O | Ensure staff have been orientated via FH education sessions on HH & its been recorded e.g.: learning hub registration. Staff have completed the Learning Hub Hand hygiene module (minimum annually) . Ensure refresh education is completed if HH audit results are under 80% | | If sites do not have learning hub access ensure documention of HH education has been recorded .There is a written hand hygiene policy and procedure that is readily available to staff. | |
| 62 | V/O | Hand hygiene results are publicly posted on each unit and in public places; results are also being shared with staff. | | Please check how they are being shared with staff e.g.: weekly meetings, visible in staff areas. AL/IL may not be aware of this requirement -ensure awareness of importance of hand hygiene practices are available in public places. | |

### COVID-19 Prevention & Outbreak Management Audit Tool For Long-Term Care/Assisted Living/Independent Living - CYCLE #2

| | | |
|---|---|---|
| **Site Name** (if floors have a significantly different population audit all floors): | | **Site Category:** |
| **Site Contact** (e.g. Residential Care Coordinator/Director of Care/Manager): | | **Site Type** |
| ***Auditor Name**: | | **Audit Type:** |
| *If auditor name is not in the list above, please enter it here: | | |
| **Audit Date (mm/dd/yyyy):** | | **Audit Time (hh:mm AM \| PM):** |

**Please Select a "Y" for Yes or "N" for No for each audit question**
**Comments (Optional)**

| # | **Legend:**<br>**1. ABHR=alcohol based hand rub**<br>**2. O=Observational/V=Verbal**<br>**3. High Risk question are identified in Red** | | **QUESTION ANSWER (Y/N)** | **TIPS for Auditors** | **Auditor's Comments**<br>*(please be as descriptive as possible )* |
|---|---|---|---|---|---|
| 63 | V/O | Behavioral Support Transition Neighbourhood (BSTN) | | Ask sites to ensure proactive individualized care plans for residents who may be at risk for non adherence to IPC measures such as : 14 day isolation, physical distancing, hand hygiene ( individual hand sani wipes) , respiratory etiquette. Encourage use of education resources: COVID-19 toolkit-refer to document for residents who cannot adhere to isolation requirements. Proactive enhanced cleaning of high touch surfaces & decluttering will reduce risk . **IF IN OUTBREAK** Site leadership may identify operational staffing needs and FH staff deployment may be utilized, ACT team may be consulted for staff support via supportive funding . **If site does not have a BSTN leave blank and comment** | |
| 64 | V/O | Pets-check if any pets in facility. In AL pets are usually in individual suites. In LTC there are roaming pets. | | Sites that have pets in the facility must have a enhanced cleaning process in common areas ( LTC) . Note in AL housekeeping is provided 1x week. Any pet must be moved to a kennell ( boarding) when in outbreak ( this includes AL) | |
| 65 | V/O | CPAP/BIPAP and Aerosoled Generating Procedures: Are their any residents/tenants on CPAP/BiPAP/nebulizers. Check N95 use. Do they have enough N95 supplies for staff who are providing direct care/monitoring. Have staff been fit tested | | **All new admits must be swabbed who require CPAP/BIPAP.** N95 is required for staff who are providing direct care for a residnet or tenant on CPAP and BIPAP & nebuilized treatment. All staff will need to be fit tested who are providing direct care/monitoring. | |
| 66 | V/O | **REGULAR AUDITS:** Hand Hygiene, PPE, Environmental cleaning ( UV glow germ audit or other product and process via Housekeeping) , Decluttering | | See section on HH , Environmental, PPE, Decluttering **.** Minimum standard is monthly/ **increase to weekly to 3 x week if outbreak, daily if complex outbreak-consult with IPC**) | |

**Other Comments**

| Long Term Care /Assisted Living / Independent Living | COVID-19 Prevention Audit Tool | |
|---|---|---|
| Site Name (if floors have a significantly different population audit all floors): | | Site Category: |
| Site Contact (e.g. Residential Care Coordinator/Director of Care/Manager): | | Site Type |
| *Auditor Name: | | Audit Type: |
| *If auditor name is not in the list above, please enter it here: | | |
| Audit Date (mm/dd/yyyy): | | Audit Time (hh:mm AM \| PM): |

| # | | Legend:<br>1. ABHR=alcohol based hand rub<br>2. O=Observational/V=Verbal<br>3. High Risk question are identified in Red | COUNT<br>YES | <br>NO | FACILITY RISK LEVEL<br>ACTION LEVEL | |
|---|---|---|---|---|---|---|
| **Entrance / Reception / Waiting Area - OBSERVATIONAL** | | | | | [NO] COUNT / PERCENT | 0% |
| 1 | VO | Alcohol-based hand rub is available & accessible & used by all who enter the facility | | | | |
| 2 | VO | Screening Process | | | | |
| 3 | VO | Signage- at entrance & exit( s) , precautions such as physical distancing, PPE, refrain from entering if ill | | | | |
| 4 | VO | Single control access point is recommended for screening and access to building * sites may have a variety of stand alone buildings so there will be more than one staff access point e.g.: campus of care. | | | | |
| 5 | VO | Ensure PPE availability eg: masks & protective eye wear are available at the entrance. | | | | |
| **Visits - Essential** | | | | | [NO] COUNT / PERCENT | 0% |
| 6 | VO | 1 essential visitor during outbreak (e.g. compassionate visits only , actively dying), visitors must be asymptomatic. | | | | |
| **Visits -Family - Social** | | | | | [NO] COUNT / PERCENT | 0% |
| 7 | V O | Family/social visits require a written plan and screening process, vistors are wearing a mask ( can be personal cloth or surgical procedural mask* ) , must be asymptomatic | | | | |
| **Personal Care Services (eg: Hairdressing, Dental Hygiene)** | | | | | [NO] COUNT / PERCENT | 0% |
| 8 | VO | Hairdressing & Personal Care Services ( written plan and site specific) | | | | |
| 9 | VO | Furniture in common areas is clean ( wipeable furniture should be cleaned twice a day( all high touch areas) . Fabric furniture should be clean, not torn and stained and cleaned q3 months | | | | |
| **Staff & Visitor Screening** | | | | | [NO] COUNT / PERCENT | 0% |
| 10 | VO | 'Active Screening' at start and end of shift (2 x shift) for all staff & visitors for: symptoms (i.e. fever, new or worsening cough, new or worsening shortness of breath, sore throat, and nausea /vomiting and diarrhea); travel outside of Canada, and/or; contact with confirmed COVID-19 case. | | | | |
| 11 | VO | All staff have been provided with information on how to self-monitor for symptoms. | | | | |
| 12 | VO | Documentation of findings of self report and checklists are kept in a binder or folder by site leaders. | | | | |
| 13 | VO | If staff are symptomatic, they are aware of process, to report to the supervisor immediately, remove themselves from work, refer to testing assessment center and self-isolate pending results. | | | | |
| **Resident/Client/Tenant Screening & Swabbing** | | | | | [NO] COUNT / PERCENT | 0% |
| 14 | VO | Active Screening' twice a day (2 x a day) as per BC CDC guidelines of all residents/persons for: symptoms (i.e. fever, new or worsening cough, new or worsening shortness of breath, sore throat, nausea /vomiting and diarrhea ); travel outside of Canada, and/or; contact with confirmed COVID-19 case. | | | | |
| 15 | VO | Is there an awareness of what to do if a resident/client/tenant becomes symptomatic, what is the process for isolating , accessing equipment and instruction for COVID-19 swabbing and notifying Public Health. | | | | |
| 16 | VO | Are site leadership aware of how to obtain nasopharyngeal ( NP) swabs and as appropriate to do NP swab & package and transport swab. Please see BCCDC guidelines and COVID toolkit. | | | | |
| **Resident/Client/Tenant Movement** | | | | | [NO] COUNT / PERCENT | 0% |
| 17 | V | Ensure awareness of transfer, admission, readmission requirements | | | | |
| 18 | O/V | Have all group activities into the community or from the community into the site stopped for LTC | | | | |
| 19 | V | Residents of LTC go into the community only for essential medical needs only . Tenants of AL & IL can engage in social and external activities aligned with general public health guidance. This may include non essential outings such obtaining essential groceries, toiletires as long as they are able to conform to the Public Health directives ( physical distancing, handwashing, respiratory etiquette) when non outbreak or prevention phase. This does not apply in an outbreak and only essential clinIcal serives and medical needs would be permitted. | | | | |
| 20 | | Resident/client/tenant may be permitted to receive essential clinical services with a onsite written plan of process | | | | |
| **Recreation** | | | | | [NO] COUNT / PERCENT | 0% |
| 21 | O/V | Congregate activities such as recreation are occurring with 2 meter physical distancing and 'no touch' practices being maintained | | | | |
| **Resident/Tenant/Client Information** | | | | | [NO] COUNT / PERCENT | 0% |
| 22 | V | All resident/persons contacts are up-to-date, including family and medical practitioners. | | | | |
| 23 | V | Does the leadership have a mechanism of contacting family with sharing of site updates or other information. | | | | |

fraserhealth
Better health. Best in health care.

| *Long Term Care /Assisted Living / Independent Living* | | | **COVID-19 Prevention Audit Tool** | | | | |
|---|---|---|---|---|---|---|---|
| | | **Site Name (if floors have a significantly different population audit all floors):** | | | **Site Category:** | | |
| | | **Site Contact (e.g. Residential Care Coordinator/Director of Care/Manager):** | | | **Site Type** | | |
| | | ***Auditor Name:** | | | **Audit Type:** | | |
| | | ***If auditor name is not in the list above, please enter it here:** | | | | | |
| | | **Audit Date (mm/dd/yyyy):** | | | **Audit Time (hh:mm AM \| PM):** | | |
| | | | **COUNT** | | **FACILITY RISK LEVEL** | | |
| # | | Legend:<br>1. ABHR=alcohol based hand rub<br>2. O=Observational/V=Verbal<br>3. High Risk question are identified in Red | **YES** | **NO** | **ACTION LEVEL** | | |
| 24 | V | All resident/persons goals of care are documented in the advance care plan and the Medical Orders Scope of Treatment ( MOST) are up-to-date (recent updating with representative/substitute decision maker). | | | | | |
| **Staffing** | | | | | **[NO] COUNT / PERCENT** | | **0%** |
| 25 | V | Is there a written staffing plan of how to obtain staff in the event of a critical shortage. | | | | | |
| 26 | V | Is there evidence that all staff have had ongoing retraining (q 3 months) on infection control ( this includes hand hygiene, PPE, and additional precautions). | | | | | |
| **Nursing Stations/ Algove/Office ( can be on multiple floors and all need to be checked) - OBSERVATIONAL** | | | | | **[NO] COUNT / PERCENT** | | **0%** |
| 27 | O | Clean, orderly and free of clutter. | | | | | |
| 28 | O | There is no food and drink. | | | | | |
| 29 | O | Clear of personal items. | | | | | |
| 30 | O | Dedicated Hand hygiene sink on unit or floor or Alcohol Based Hand Rub (ABHR) is available & accessible (e.g. container is not empty). | | | | | |
| **Soiled Utility Room/Housekeeping Room/Laundry Room** | | | | | **[NO] COUNT / PERCENT** | | **0%** |
| 31 | O | Door to room is kept closed and restricted access. | | | | | |
| 32 | O | Hand hygiene sinks have paper towel and plain liquid soap dispensers in close proximity (e.g. ensure container is not empty). | | | | | |
| 33 | O | Use of precautions when doing Laundry and observations in Laundry room as a common area (tips apply particularly in IL as a campus of care). | | | | | |
| 34 | O | The hand hygiene sink is completely free from surrounding clutter. | | | | | |
| **Clean and Sterile Supply Room** | | | | | **[NO] COUNT / PERCENT** | | **0%** |
| 35 | V | Sterile storage has a designated area and is limited to only staff access and door to room is kept closed. | | | | | |
| 36 | V&O | PPE are kept in secured area. | | | | | |
| **Hallway - Observational** | | | | | **[NO] COUNT / PERCENT** | | **0%** |
| 37 | O | ABHR is available and accessible outside residents/tenants room or common area in AL | | | | | |
| 38 | O | Free of clutter- carts, wheelchairs, equipment. | | | | | |
| **Dining areas e.g. kitchenettes, servery, eating area** | | | | | **[NO] COUNT / PERCENT** | | **0%** |
| 39 | O | Kitchenettes are closed. | | | | | |
| 40 | O | Ensure scheduled cleaning of all surfaces in dining area after every meal, following every sitting, minimum of twice a day. | | | | | |
| 41 | O | Environmental controls are in place to ensure physical distancing (2 metre distancing). | | | | | |
| 42 | O | Hand Hygiene sink/ABHR is available (e.g. container is not empty). | | | | | |
| 43 | V | There is a process for cleaning residents/tenants hands before/after meal time. | | | | | |
| 44 | O | There is signage indicating step-by-step guide to proper hand hygiene (HH). | | | | | |
| 45 | O/V | Furniture cleanable and able to withstand disinfection with hospital grade disinfectant. | | | | | |
| **Housekeeping** | | | | | **[NO] COUNT / PERCENT** | | **0%** |
| 46 | O/V | Is enhanced cleaning in place for the entire facility, with high touch surfaces cleaned and disinfected twice a day (6-8 hours between cleaning). | | | | | |
| 47 | O/V | Regularly scheduled cleaning of housekeeping equipment e.g.: mop heads; toilet brushes, according to protocol. | | | | | |
| 48 | | Garbage bins are recommended to be handsfree in all areas | | | | | |
| 49 | | Staff rooms: Cleaning between use, after use, between shifts, ensuring environmental controls for physical distancing , scheduled and staggered breaks. | | | | | |
| 50 | O | Sites demonstrate and show what they are using for cleaning -should be .5% accelerated hydrogen peroxide or hospital grade disinfectant eg: bleach wipes/cavi wipes. Health Canada approved disinfectants for COVID-19. | | | | | |
| **Reprocessing -Cleaning instructions** | | | | | **[NO] COUNT / PERCENT** | | **0%** |
| 51 | O/V | Cleaning instructions ( work) are available for all shared equipment and recreational equipment (e.g. lifts, walkers, wheelchairs, slings both sit to stand and ceiling lifts, shower chair, tub; recreation tools such as bowling equipment, instruments) | | | | | |
| 52 | O/V | There is no sharing of equipment , wash basins or supplies between residents/persons without appropriate cleaning and disinfection between use. | | | | | |
| **PPE** | | | | | **[NO] COUNT / PERCENT** | | **0%** |
| 53 | V | PPE is available, stored appropriately and accessible for all staff. Staff are also provided what is required for direct care to residents. | | | | | |

fraserhealth
Better health. Best in health care.

| *Long Term Care /Assisted Living / Independent Living* | | | **COVID-19 Prevention Audit Tool** | | | |
|---|---|---|---|---|---|---|
| **Site Name (if floors have a significantly different population audit all floors):** | | | | | | **Site Category:** |
| **Site Contact (e.g. Residential Care Coordinator/Director of Care/Manager):** | | | | | | **Site Type** |
| ***Auditor Name:** | | | | | | **Audit Type:** |
| ***If auditor name is not in the list above, please enter it here:** | | | | | | |
| **Audit Date (mm/dd/yyyy):** | | | | | | **Audit Time (hh:mm AM \| PM):** |
| | | | **COUNT** | | **FACILITY RISK LEVEL** | |
| # | Legend:<br>1. ABHR=alcohol based hand rub<br>2. O=Observational/V=Verbal<br>3. High Risk question are identified in Red | | **YES** | **NO** | **ACTION LEVEL** | |
| 54 | O/V | Is a three day supply of PPE available on site (including surgical/procedural masks, gloves, gowns, and eye protection)? | | | | |
| 55 | O | Staff are wearing recommended PPE when in resident common areas & when providing direct care. The recommended PPE must be worn at all times in these circumstances. They are surgical/procedural mask and protective eye wear. | | | | |
